# Potential role of aberrant mucosal immune response to SARS-CoV-2 in pathogenesis of IgA Nephropathy

**DOI:** 10.1101/2020.12.11.20247668

**Authors:** Zhao Zhang, Guorong Zhang, Meng Guo, Wanyin Tao, Xing-Zi Liu, Haiming Wei, Tengchuan Jin, Yue-Miao Zhang, Shu Zhu

**Author notes:** Equal contribution (Zhao Zhang, Guorong Zhang, Meng Guo). Corresponding author: Professor Shu Zhu, Or Dr Yue-miao Zhang.

## Abstract

Aberrant mucosal immunity has been suggested to play a pivotal role in pathogenesis of IgA nephropathy (IgAN), the most common form of glomerulonephritis worldwide. The outbreak of severe acute respiratory syndrome coronavirus (SARS-CoV-2), the causal pathogen of coronavirus disease 2019 (COVID-19), has become a global concern. However, whether the mucosal immune response caused by SARS-CoV-2 influences the clinical manifestations of IgAN patients remains unknown. Here we tracked the SARS-CoV-2 anti-receptor binding domain (RBD) antibody levels in a cohort of 88 COVID-19 patients. We found that 52.3% of the COVID-19 patients produced more SARS-CoV-2 anti-RBD IgA than IgG or IgM, and the levels of the IgA were stable during 4-41 days of infection. Among these IgA-dominated COVID-19 patients, we found a severe COVID-19 patient concurrent with IgAN. The renal function of the patient declined presenting with increased serum creatinine during the infection and till 7 months post infection. This patient predominantly produced anti-RBD IgA as well as total IgA in the serum compared to that of healthy controls. The analysis of the IgA-coated microbiota as well as proinflammatory cytokine IL-18, which was mainly produced in the intestine, reveals intestinal inflammation, although no obvious gastrointestinal symptom was reported. The mucosal immune responses in the lung are not evaluated due to the lack of samples from respiratory tract. Collectively, our work highlights the potential adverse effect of the mucosal immune response towards SARS-CoV-2, and additional care should be taken for COVID-19 patients with chronic diseases like IgAN.

## Introduction

IgA nephropathy (IgAN) is the most common form of glomerulonephritis worldwide [1-3], characterized by deposition of IgA or IgA-containing circulating immune complexes in glomerular mesangium [3, 4]. Although the cause of IgAN remains unclear, IgA was suggested to play a key role in the disease pathogenesis. Evidence from genetic, experimental and clinical studies has suggested the involvements of genetic predisposition and infection [5]. Etiologic factors may trigger an aberrant mucosal immune IgA response either in lung, or in intestine, interacting with genetic predisposition, leading to the onset and progression of IgAN [5-8]. For example, seasonal flu infections were reported to trigger the pathogenesis of IgAN [9-11]. Currently, the highly contagious, rapidly spreading SARS-CoV-2 has caused a global pandemic. However, whether and how the mucosal immune response caused by SARS-CoV-2 influences the progression of IgAN remains unknown.

The angiotensin-converting enzyme 2 (ACE2) is the receptor required for cellular entry of SARS-CoV-2 [12, 13], which was highly expressed in both lung and intestine [14, 15]. Consistently, although pneumonia is the most common symptom in patients with moderate to severe illness [16-21], 17.6% of COVID-19 patients developed gastrointestinal symptoms, including diarrhea, anorexia and nausea [22-25]. It was reported that 12 out of 173 patients who recovered from COVID-19 were re-detectable positive in SARS-CoV-2 RNA test, which was found to be associated with the potential intestinal infection of SARS-CoV-2 in these patients [26-29]. Moreover, besides direct gastrointestinal infection, recent studies reported SARS-CoV-2 results in intestinal dysbiosis of the microbiota, along with the increase of the opportunistic pathogens in the intestine [30-32].

IgA is the most abundant antibody isotype in the mucosal immune system such as intestine and lung to offer humoral protection against microbial pathogens [33]. Ejemel et al. showed human anti-SARS-CoV-2 IgA efficiently neutralize the virus at mucosal surface by binding to the spike protein of SARS-CoV-2 [34]. Several studies also demonstrated the role of serum anti-SAR-CoV-2 IgA in diagnosing of recovered COVID-19 patients [34-38]. However, mucosal IgA responses may also promote the disease pathogenesis of IgA vasculitis with nephritis (Henoch-Schönlein purpura) and Kawasaki disease (KD) [39-42]. Thus, this study was designed to systemically evaluate the mucosal humoral immune response towards SARS-CoV-2 and its potential role in disease progress of IgAN.

Intriguingly, we found more than half of COVID-19 patients showed an IgA-dominant phenotype, suggests that SARS-CoV-2 induced strong mucosal immune response. One of the IgA-dominant patients, who was diagnosed with IgAN before, was found to get a declined renal function during and after recovered from SARS-CoV-2 infection, due to high levels of IgA in both serum and feces. Besides the severe pneumonia, this patient also produced higher pro-inflammatory cytokine from the intestine. Thus, we speculate that mucosal immune responses towards SARS-CoV-2 in intestine and lung may worsen the renal function and promote the IgAN progression in IgAN patients. Extra care should be taken for COVID-19 patients with chronic diseases like IgAN.

## Material and Method

### Patient cohort

88 COVID-19 patients with average age of 47.35±15.69 years (range 21-91) were enrolled, including 88 patients from the First Affiliated Hospital of USTC and the First Affiliated Hospital of Anhui Medical University [43] and one patient concurrent with IgAN who underwent kidney transplant (hereafter referred as COVID-19 IgAN case; Supplementary Note 1) from People’s Hospital of Wuhan University. Specifically, all patients had positive testing for viral nucleic acid of SARS-CoV-2 (Real-Time Fluorescent RT-PCR Kit, BGI, Shenzhen). Among them, six were critically ill and admitted to intensive care unit, one of which was died of cerebral hemorrhage after stroke. Seventeen patients were severely infected and all of them received oxygen supplementation treatment. Fifty-six patients showed moderate illness and nine patients showed mild infection. Underline symptoms were found in thirty-seven (42.0%) patients; hypertension in eighteen (20.5%) was the most common one.

A total of 218 serum samples collected during the hospitalization and after discharge were tested for SARS-CoV-2 spike (S) protein specific antibodies. 31 (35.2%), 19 (21.6%), 16 (18.2%), 12 (13.6%), 8 (9.1%), 1 (1.1%), and 1 (1.1%) patients’ blood were collected for 1, 2, 3, 4, 5, 6, and 7 times, respectively. This cohort contains 50 archived sera from healthy donors collected before October 2019 as healthy controls to evaluate the reliability of the measurements. Besides, urine and fecal samples from the COVID-19 IgAN case at 5-month (day 160) and 7-month (day 209) after discharge were also collected. Serum, urine and fecal samples from 5 matched healthy controls were correspondingly collected for the COVID-19 IgAN case.

This study was reviewed and approved by the Medical Ethical Committee of the First Affiliated Hospital of USTC (approval number: 2020-XG(H)-014) and the First Affiliated Hospital of Anhui Medical University (approval number: Quick-PJ 2020-04-16).

### Measurement of serum immunoglobins

SARS-CoV-2 specific IgA, IgM and IgG detection kits using chemiluminescent method were developed by Kangrun Biotech (Guangzhou, China), in which the receptor binding domain of spike was coated onto magnetic particles to catch SARS-CoV-2 specific IgA, IgM and IgG in patient samples. A second antibody that recognizes IgA, IgM, or IgG conjugated with acridinium (which can react with substrates to generate a strong chemiluminescence) was used for detecting IgA, IgM and IgG, respectively. The detected chemiluminescent signal over background signal was calculated as relative light units (RLU). It has been validated in a large cohort of serum samples showing high sensitivities and specificities [44]. Serum samples were collected by centrifugation of whole blood in test tubes at room temperature for 15 min, 1000 x g. Prior to testing, a denaturant solution was added to each serum to a final concentration of 1% TNBP, 1% Triton X-100. After adequate mixing by inverting, the samples were incubated at 30°C for 4 hours to completely denature any potential viruses. Virus-inactivated serum samples were then diluted 40 times with dilution buffer and subjected to testing at room temperature. Then RLU was measured using a fully automatic chemical luminescent immunoanalyzer, Kaeser 1000 (Kangrun Biotech, Guangzhou, China).

### Measurement of total IgA and IL-18

For the COVID-19 IgAN patient, total IgA and IL18 were also detected. The total IgA in the serum, urine and fecal was measured using a human Immunoglobulin A (IgA) ELISA kit (CUSABIO, CSB-E07985h) according to manufacturer’s instruction. The serum IL-18 was detected by ELISA Kit (Sino Biological) according to instructions of the vendor.

### Fecal IgA flow cytometry

Fecal samples of the COVID-19 IgAN patient were stored at −80 °C until the time of following analyses. Fecal pellets collected directly from frozen human fecal material were placed in Fast Prep Lysing Matrix D tubes containing ceramic beads (MP Biomedicals) and incubated in 1 mL Phosphate Buffered Saline (PBS) per 100 mg fecal material on ice for 15 min. Fecal pellets were homogenized by bead beating for 5 seconds and then centrifuged (50 x g, 15 min, 4°C) to remove large particles. Fecal bacteria in the supernatants were removed (100 µL/sample), washed with 1 mL PBS containing 1% (w/v) Bovine Serum Albumin (BSA, American Bioanalytical; staining buffer) and centrifuged for 5 min (8,000 x g, 4°C). After an additional wash, bacterial pellets were resuspended in 100 µL blocking buffer (staining buffer containing 20% Normal Mouse Serum for human samples, from Jackson Immuno Research), incubated for 20 min on ice, and then stained with 100 µL staining buffer containing PE-conjugated Anti-Human IgA (1:10; Miltenyi Biotec clone IS11-8E10) for 30 minutes on ice. Samples were then washed 3 times with 1 mL staining buffer before flow cytometric analysis.

### Statistical analysis

The sample size chosen for our experiments in this study was estimated based on our prior experience of performing similar sets of experiments. For all the bar graphs, data were expressed as mean ± SEM. A standard two-tailed unpaired Student’s t-test was performed using GraphPad Prism 7. P values ≤ 0.05 were considered significant. The sample sizes (biological replicates), specific statistical tests used, and the main effects of our statistical analyses for each experiment were detailed in each figure legend.

## Results

### More than half of COVID-19 patients produced more anti-RBD IgA than IgG or IgM during SARS-CoV-2 infection

Analysis of the SARS-CoV-2 anti-RBD antibodies in the serum of the 88 COVID-19 patients showed that 46 (52.3%) patients were IgA-dominated during the infection (Figure 1a). The IgA level was the highest in IgA-dominated group comparing to those in healthy controls (1378338±198038, n=46 vs. 10424±747.3, n=50, *P<*1*10^−4^), IgG-(480603±57131, n=28, *P=*0.0009), and IgM-dominated (380694±89277, n=14, *P=*0.0082) group (Figure 1b). Notably, the IgA level are largely stable within 4 to 41 days since the disease onset (Figure 1c).

**Figure 1.**
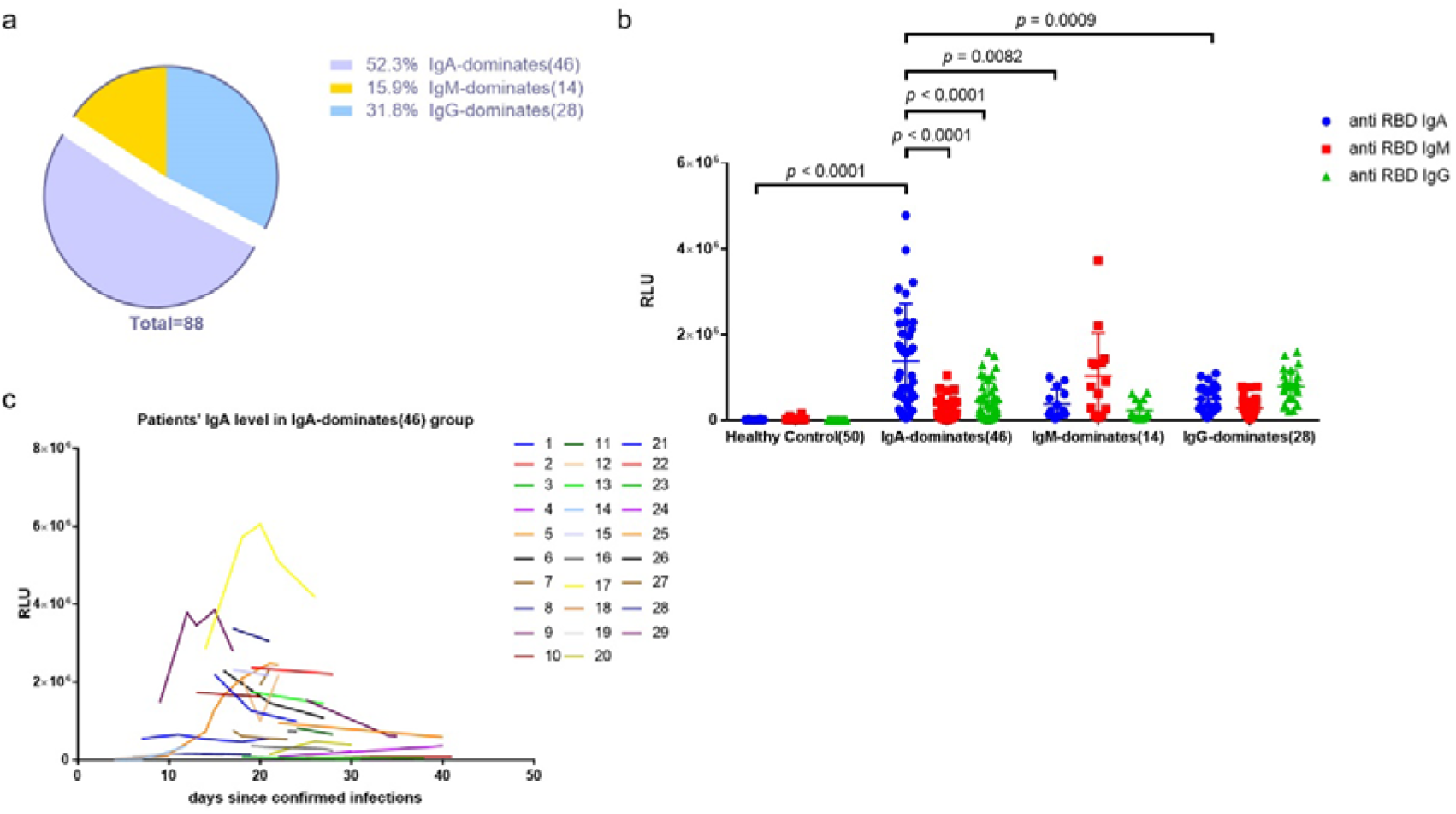
Analysis of the anti-RBD antibodies of a cohort of 88 patients during the infection. **a**. percentage of IgA-, IgM-, and IgG-dominated patients in the cohort of COVID-19 patients (n=88); **b**. Levels of anti-RBD-IgA, IgM, IgG in 88 COVID-19 patients and 50 healthy controls. RLU: relative light unit. Statistical significance was determined using two-tailed Unpaired Student’s *t* test. **c**. Duration of anti-RBD-IgA levels in IgA-dominant COVID-19 patients.

### An IgA-dominant COVID-19 patient concurrent with IgAN showed declined renal function during and post infection

We found one IgA-dominant COVID-19 case, who had a history of IgAN and underwent kidney transplantation 25 months before the infection. Prednisone (5 mg/d), tacrolimus (5 mg/d), and mycophenolate (0.5 g/d) were applied for post-surgery treatment. The post-surgery urinary protein was around 0.15 g/L and serum creatinine was around 160 μmol/L.

On 16^th^ Jan 2020, the patient was hospitalized due to symptoms including low fever of 37.4 °C, fatigue and dry cough. No gastrointestinal symptom such as nausea, vomiting, and diarrhea, was reported. Primary microbial tests, including Influenza A virus, influenza B virus, parainfluenza virus, respiratory syncytial virus, metapneumovirus, coronavirus, rhinovirus, adenovirus, Boca virus, and mycoplasma pneumoniae virus, were all negative. Computer tomography (CT) of chest showed infectious lesions in both lungs. During 16^th^ Jan to 20^th^ Jan, her body temperature fluctuated between 36.9 and 38 °C. White blood cells (4.24×10^9^/L, reference: 3.5∼9.5×10^9^/L) was within the normal range; Lymphocyte count was low (0.60×10^9^/L, reference: 1.0∼3.2×10^9^/L); CD4+ T cell count was 186×10^9^/L (reference: 404∼1,612×10^9^/L). On 20^th^ Jan (day 1), a positive reverse transcriptase-polymerase chain reaction (RT-PCR) assay via nasopharyngeal swab confirmed SARS-CoV-2 infection. During the infection period, serum creatinine of the patient increased to 197 μmol/L. Worse proteinuria was also reported but 24-hour urine protein was not measured due to medical limitations at the very moment. The patient condition deteriorated into respiratory failure and required ventilatory support. Specific COVID-19 management with immunosuppression reduction (mycophenolate withdrawal and tacrolimus withdrawal) was attempted immediately, including methylprednisolone (40 mg/d) injected intravenously for anti-inflammation, human blood gamma globulin (10 g/d) injected intravenously for immunity enhancement, moxifloxacin hydrochloride (4.5 g/d) and sulfanilamide (6 pieces/d) for anti-microbial infection, Posaconazole (30 ml/d) for anti-fungi infection, and Aciclovir (250 mg/d)/Oseltamivir (150 mg/d) for antiviral treatment. The patient’s condition was obviously improved and stable afterwards. However, the serum creatinine levels went to the top (208 μmol/L) at 4 months and remained high (190-195 μmol/L) even after 7 months post infection. Clinical and laboratory characteristics are shown in Table 1, Figure 2.

**Table 1.** The clinical and laboratory characteristics of the COVID-19 IgAN case.

| **Clinical Characteristics** | **COVID-19 IgAN case** |
| --- | --- |
| Age, y | 39 |
| Sex | Female |
| Cause of kidney failure | IgAN |
| Kidney failure vintage, y | 14 |
| Signs and symptoms of SARS-CoV-2 infection |  |
| Fever | Yes |
| Dry cough | Yes |
| dyspnea | Yes |
| Fatigue | Yes |
| Nausea | No |
| Vomiting | No |
| Diarrhea | No |
| Gross hematuria | No |
| **Laboratory Characteristics** |  |
| White blood cell count, $\times 10^3/\mu\text{L}$ | 4.24 |
| Neutrophil count, $\times 10^3/\mu\text{L}$ | 3.16 |
| Lymphocyte count, $\times 10^3/\mu\text{L}$ | 0.6 |

**Figure 2.**
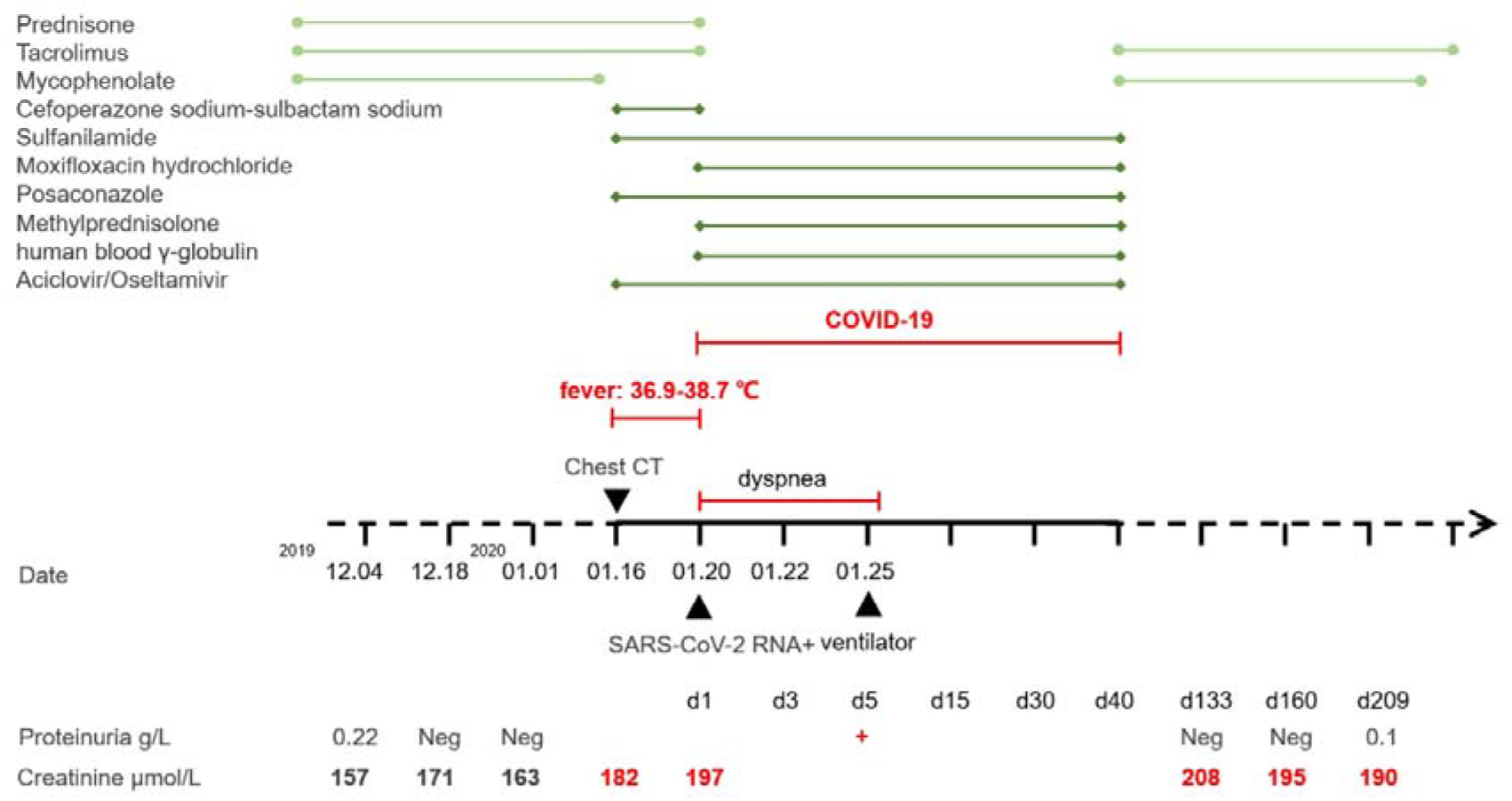
The clinical manifestations, medications and parameters of renal functions of the COVID-19 IgAN case.

### Intestinal inflammation was observed in the COVID-19 IgAN case

We first tracked the SARS-CoV-2 anti-RBD IgA, IgG, IgM antibodies in the serum, urine, and feces of this COVID-19 IgAN case and compared with those from healthy controls. Results revealed that all the SARS-CoV-2-specific IgA, IgG and IgM responses were higher in the serum of the case compared to healthy controls, with the IgA increase being the most significant (Figure 3a). To our surprise, the IgA antibody retained high even though it was 7 months post infection (anti-RBD IgA RLU: 138475±26834, anti-RBD IgM RLU: 18084±967, anti-RBD IgG RLU: 36991±7665). However, the increase of anti-RBD IgA, IgG, IgM antibodies was not observed in urine or fecal samples of this COVID-19 IgAN case. (Figure 3a). The anti-RBD IgA, IgG, IgM levels in respiratory samples and mucosal samples in both intestine and lung were not measured due to the limitations to collect these samples.

**Figure 3.**
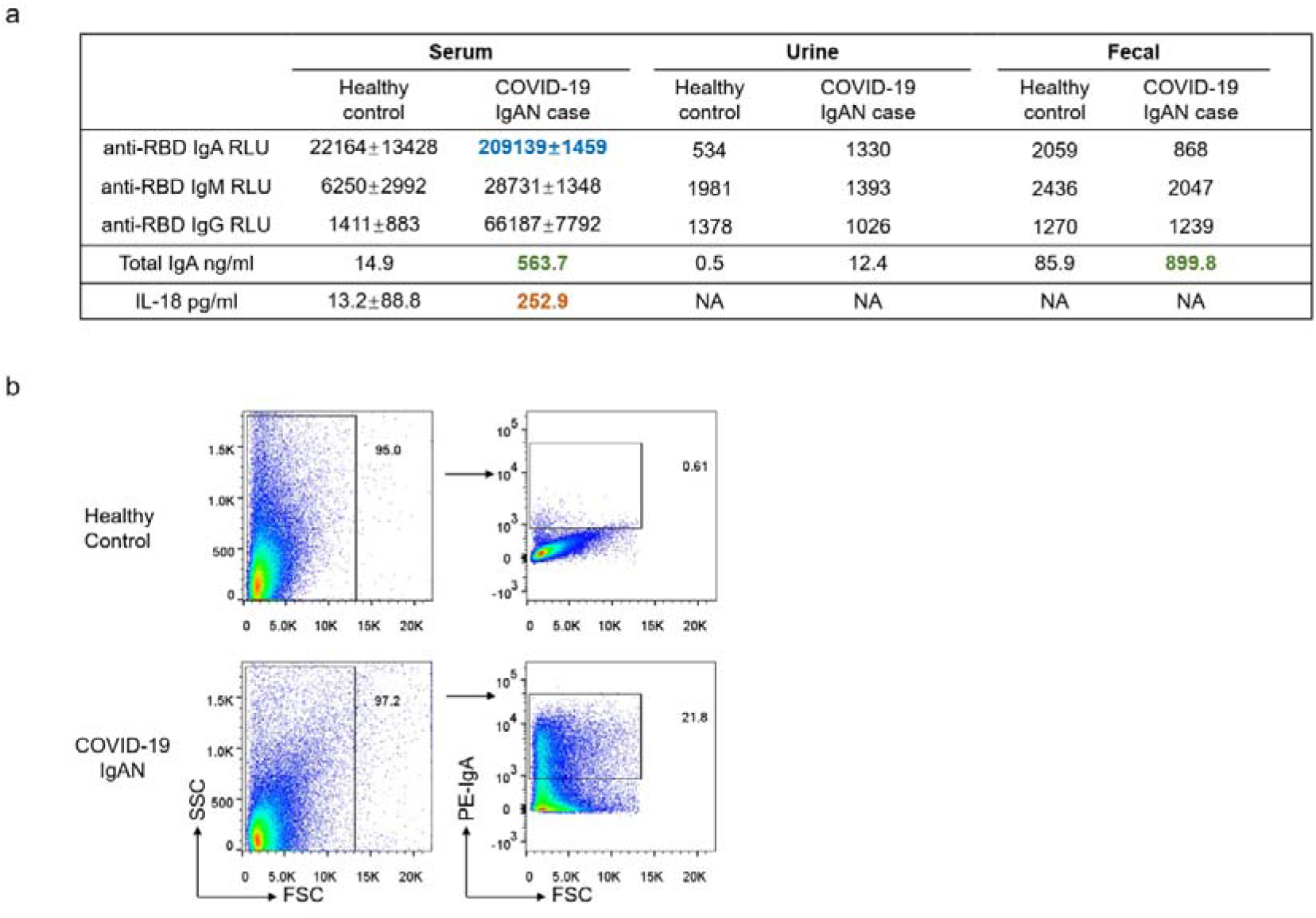
Measurements of mucosal immune responses and microbiota in the COVID-19 IgAN case.

Interestingly, total IgA was much higher in both serum and fecal samples of this patient compared to that of healthy control (Figure 3a), leading to the hypothesis that the infection in the intestine may contribute to the production of IgA. Therefore, accompanied by increased IgA-coated microbiota (Figure 3b), higher serum IL-18, a primary mediator of the inflammation [45], also reflects a more inflamed gut (Figure 3a).

## Discussion

Herein, we observed that SARS-CoV-2 as an infectious agent activated aberrant mucosal humoral response both in lung and intestine, which may have a long-term side-effect on renal function in an IgAN patient recovered from COVID-19. By focusing on the humoral response towards SARS-CoV-2 in a cohort of COVID-19 patients, we found that the SARS-CoV-2 anti-RBD IgA was the leading functional isotype in the patients’ serum during the infection and the ratio of patients presented with IgA-dominant response is as high as 52.3%. Further, among these COVID-19 patients with IgA-dominant humoral response, we found a patient who had a history of IgAN showed severe pneumonia. An increased serum creatinine and worse proteinuria were observed during and even after the patient recovered from the SARS-CoV-2 infection. In addition to severe pneumonia, the patient developed intestinal dysbiosis and produced higher pro-inflammatory cytokine from the intestine. Collectively, our work revealed that aberrant mucosal IgA against SARS-CoV-2 may contribute to kidney pathogenesis and IgAN progression.

The hit1 of the classic “Four Hits theory” of IgAN pathogenesis begins with IgA production and abnormalities in circulating IgA [46]. The response of the mesangium and the deposited IgA is crucial to the development of IgAN [47]. The source of these pathogenic IgA in IgAN has been an emerging area of study. Both mesangial IgA and the increased fraction in serum IgA are polymeric, which is normally produced at mucosal surfaces, leading to the suspicion that IgAN was intimately linked with abnormal mucosal immune response against microorganisms [47]. For example, episodic macroscopic hematuria after upper respiratory infections was common in IgAN patients [48]. Tonsillectomy can improve urinary findings and IgA deposits, thus have a favorable effect on long-term renal survival in some IgAN patients [49, 50]. Since IgA is widely found in the gut mucosal immune system, gut microbiota dysbiosis has also been found to play a role in the pathogenesis of IgAN [51-53]. Exclusive differences in gut microbiota composition has been investigated in patients with IgAN and healthy controls [52]. Targeted release of budesonide to the distal ileum was reported to reduces proteinuria and stabilize renal function in IgAN patients [54]. In addition, recent studies reported that the gut microbiota signature of COVID-19 patients was different from that of healthy controls [31]; COVID-19 patients showed a significant dysbiosis of fecal microbiome, characterized by enrichment of opportunistic pathogens and depletion of beneficial commensals [30]. Results from our experiments showed that total IgA was also much higher in the serum and fecal of this patient compared to that of healthy control; the gut microbiota dysbiosis and higher level of proinflammatory cytokine were also identified in this patient.

Mucosal humoral response protects human from viral infections. IgA responses were detected in patients infected with influenza A(H1N1) pdm09 virus and a >4-fold increases in IgA response to A(H1N1) pdm09 hemagglutinin (HA) was found in 67% of A(H1N1) pdm09-infected persons [55]. In the current pandemic, research found that SARS-CoV-2-specific IgA dominates the early neutralizing antibody response against SRAS-CoV-2 [56]. The IgA level was higher in critically ill patients compared to those with mild to moderate illness [43, 57]; and it peaked at the first to third weeks of onset and declines after one month [43, 56]. Based on an estimated decline rates of virus-specific antibodies using a previously established exponential decay model of antibody kinetics after infection, researchers reported that the predicted days when convalescent patients’ anti-RBD IgA reaches to an undetectable level are approximately 108 days after hospital discharge [58]. A preprint by Gaebler et al. reports that in a longitudinal analysis of SARS-CoV-2-specific humoral responses in 87 patients, memory B cells potentially generating anti-RBD IgA persisted up to 6 months after infection and displayed with increased neutralization potency and breadth [59]. Further, study reported peripheral expansion of mucosal-homing IgA-plasmablasts cells at the early onset of the symptom and peaked during third week of the infection [56]. Reliable evidence has been provided in supporting that the SARS-CoV-2 anti-RBD IgA titer in the serum would accurately reflect that of anti-SARS-CoV-2 sIgA [60, 61]. These evidences suggested that assessment of serum SARS-CoV-2 anti-RBD IgA can yield reliable information of anti-SARS-CoV-2 mucosal immunity. In the current study, the COVID-19 IgAN patient produced predominately anti-RBD IgA and the level was still high until month 7, during this time the kidney function was found to be reversely associated with serum IgA level. Therefore, the mucosal immune response against SARS-CoV-2 might have contributed to the progression of IgAN in this patient.

Nearly one year into the COVID-19 pandemic, which has infected nearly 69,000,000 people globally and caused over 1,600,000 deaths by December, 2020, there is still much that we do not understand. However, we shall be better prepared in handling the COVID-19 patients comparing to that in the initial unexpected exposure. The impact of COVID-19 on glomerular disease has been largely contracted to acute kidney injury, which occurred in nearly 46% of hospitalized patients [62]. Like the majority of kidney diseases, the mechanisms are mostly likely the direct viral infection, inflammatory syndrome-mediated injury, hemodynamic instability and the hypercoagulable state, all of which may have happened during infection. However, presentation of one case reporting concurrent IgAN after SARS-CoV-2 recovery raised great concerns regarding to the hidden and prolonged impact SARS-CoV-2 infection may have caused [63]. Most concerning is the persistence of IgA antibody and memory B cells with the IgA-generating potency in patients who have already recovered from COVID-19. We have yet to fully appreciate their potential for progression to IgAN or even end-stage kidney disease. Thus, additional care should be taken for especially COVID-19 IgAN patients. For example, a complete medical history needs to be taken for newly diagnosed COVID-19 patients to alert doctors with additional aspects of disease progression. Further, like the COVID-19 IgAN case we reported here, others who recovered from the infection and are prone to develop IgA-related disease are suggested to complete a regular follow-up, so as to monitor disease conditions properly. Least but not least, it is important to avoid all forms of infections that might result in IgA abnormalities.

We acknowledge that the limitation of the study is the shortness of patient and a lack of evidence from kidney biopsy in the only COVID-19 IgAN case. Nonetheless, the long-term regular follow-up, strict supervision of the patient’s kidney condition, and our laboratory evidence suggest that the high level of mucosal IgA response to SARS-CoV-2 is closely associated with the declined renal function.

In conclusion, our study indicates that SARS-CoV-2 infection stimulates aberrant mucosal humoral IgA in the lung and intestine against SARS-CoV-2 and may have contributed to kidney damage and potentially promoted IgAN progression. Thus, our work highlights the potential adverse effects of the humoral immune response towards SARS-CoV-2, and additional care should be taken for COVID-19 patients with chronic diseases like IgAN.

## Data Availability

The data that support the findings of this study are available from the corresponding author, [S.Z.], upon reasonable request.

## FOUNDING

This work was supported by a grant from the Strategic Priority Research Program of the Chinese Academy of Sciences (XDB29030101) (SZ), National Key R&D Program of China (2018YFA0508000) (SZ), and National Natural Science Foundation of China (81822021, 91842105, 31770990, 81821001) (SZ).

## ACKNOWLEDGMENTS

We thank Dr. Tian ZG, Dr. Zhou RB, and Dr. Weng JP for discussion and comments.

## SUPPLEMENTAL MATERIAL

Supplementary Note 1.

The COVID-19 IgAN case is a female, 39-yesr-old. She developed proteinuria 19 years ago, taking traditional Chinese medicine for treatment, and the urinary protein was around 2.0 g/L; no kidney biopsy was performed. About 14 years ago, she observed massive proteinuria, and urinary protein was increased to 5.0 g/L, kidney biopsy was performed. Pathological diagnosis indicated IgAN, including diffused mesangial hyperplasia with focal nodular sclerosis and mild renal tubulointerstitial lesions. She underwent kidney transplantation 25 months ago. Prednisone (5 mg/d), tacrolimus (5 mg/d), and mycophenolate (0.5 g/d) were applied for after surgery treatment. Urinary protein reduced to 0.22 g/L and 0.15 g/L, 22.5 months and 24 months after surgery, respectively. Serum creatinine was around 170 μmol/L. No other abnormal indicators were reported.

